# Painting observation changes balance in patients with bilateral vestibulopathy

**DOI:** 10.1101/2025.05.20.25327995

**Authors:** Adéla Kola, Gaëlle Quarck, Olga Kuldavletova, Thomas Stoffregen, Pierre Denise, Antoine Langeard

## Abstract

**Background:** Bilateral vestibulopathy is a disorder characterized by significant impairments in vestibular function, leading to changes in the kinematics of standing body sway. While previous studies have demonstrated that observing paintings can influence postural control in healthy adults, the effects of such visual stimuli on individuals with BVP remain unexplored. This study aimed to investigate the impact of observing an artistic painting on postural balance.

**Methods:** Posture was assessed in 34 patients with bilateral vestibuloapthy compared to 30 healthy controls using three static balance conditions: (1) eyes open by fixating a cross, (2) eyes closed, and (3) while viewing an impressionist painting (Le Bassin Aux Nymphéas, Monet).

**Results:** Patients with bilateral vestibulopathy, compared to healthy controls, had higher center of pressure standard deviation and amplitude in the anteroposterior direction eyes open and eyes closed but no differences between groups were detected when viewing the painting.

**Conclusions:** These findings suggest that art observation can influence postural control in vestibular-defective patients. Further research will be needed to understand the basis of this effect and its possible relevance for rehabilitative treatment.

## Introduction

Vestibular disorders are associated with changes in postural activity. Bilateral vestibulopathy (BVP) is a rare pathology affecting approximately 28 cases per 100,000 individuals in 2008 in the United States (1). This rare disorder is often idiopathic and, in some cases, may be related to ototoxicity induced by pharmacological agents such as gentamicin (2). This disorder leads to increased body sway (1). For patients with BVP, sway becomes more pronounced during stance on irregular or soft surfaces, and in low-light conditions (2). Similarly, eye closure is associated with significant increases in sway in both the mediolateral and anterior-posterior axes (3), highlighting the role of vision in postural control for these patients. However, the existing literature provides little insight into the control of standing posture among BVP patients in relation to naturalistic variations in visual activity.

Some studies have revealed that postural control is modulated according to the demands of visual tasks. For example, compared with looking a blank environment, reading a text reduces the magnitude of sway (4,5). This effect persists in healthy elderly individuals (6,7), and in some clinical populations (8,9). By contrast, adults with dementia do not modulate their sway in response to variations in visual tasks (10). Similar effects of visual tasks on postural activity have been observed in relation to tasks that were based on visual graphics, rather than text (8). In the present study, we investigated whether patients with BVP would modulate their standing body sway in relation to viewing visual graphics in the form of artistic paintings.

Artistic paintings, which evoke psychological states (11) and neural responses (12), hold promise for improving our understanding of the complex interaction between vision and postural balance in patients with BVP. In healthy individuals, the kinematics of standing body sway can be influenced by viewing of both figurative (13) and abstract art (14,15). We asked whether effects that have been observed in healthy populations would also be found in patients with BVP. Specifically, we examined the effects of looking at an impressionist painting on postural stability in individuals with BVP compared to healthy control in comparison with standard postural assessment. By comparing postural control under different visual conditions (eyes open, eyes closed, and while viewing a painting), we investigated whether the postural responses of patients with BVP would be similar to those of healthy controls.

## Methods

### Participants

The study included 34 patients diagnosed with BVP and 30 healthy control participants (see Table 1). Ethical approval was obtained from the French Ethical Committee (Comité de Protection des Personnes de la Région Ouest I, ID-RCB 2022-AO1513-40). BVP subjects were members of the *Association Française de Vestibulopathie Bilatérale* (www.afvbi.info). The BVP diagnosis followed the Bárány Society criteria, as described by Strupp et al. (2017) (1). All participants were tested in the COMETE Laboratory at the University of Caen from 3 October 2022 to 18 January 2024.

**Table 1.** Subject’s characteristics (mean+/-sd)

| Characteristics | BVP | Controls |
| --- | --- | --- |
| Sex | 19 ♀; 15 ♂ | 17 ♀; 13 ♂ |
| Age (years) | 60 ± 13 | 59 ± 13 |
| Education level (years) | 2.76 ± 2.55 | 2.83 ± 2.13 |
| Height (cm) | 169.6 ± 8.42 | 168.3 ± 9.16 |
| Foot length (mm) | 250.73 ± 16.59 | 252.90 ± 16.54 |

### Experimental protocol

Postural control was assessed using the Synapsys Posturography System® (SPS; Synapsys, Marseille, France; sampling frequency of 100 Hz). Safety rails were installed around the platform to ensure participants’ safety and to prevent falls. Participants stood barefoot on a force platform, with their feet positioned at a 30° angle (16), and were instructed to keep their arms relaxed at their sides without lifting their feet.

Participants performed 60-second tasks in three different visual conditions: 1) eyes open (EO), where participants fixated a cross positioned at eye level in front of them; 2) painting exploration, where participants visually explored an impressionist painting (*Le Bassin aux nymphéas, Monet*); and 3) eyes closed (EC), were participants stood with their eyes closed. The tests were performed in a random order. The distance between the subject and the image (size: 172x113.5 cm) was 190 centimeters. We excluded from analysis trials on which the participant had any physical contact with the safety rails.

The SPS system calculated measures of postural balance, including amplitudes (ranges of maximum-minimum of CoP displacement) (CoP-amplitude), velocities (CoP-velocity), and standard deviations (CoP-SD) in the mediolateral (ML) and anteroposterior (AP) directions.

### Statistical analyses

Robust linear mixed models were used to compare the group and task differences, controlling for sex, age, education level, height, and foot length. When a Group × Condition interaction was significant, the Tukey method was used for multiple comparison corrections in post hoc analyses. The significance threshold was set at a p-value of 0.05. The Group × Condition were calculated with the painting condition as a reference. The statistical analyses were conducted in the R Studio Software (2023.12.1+402), using the rlmm package.

## Results

The data and results are summarized in Table 2 and in Supplementary Table.

**Table 2.** Marginal means (±SE) by groups (controls and BVP patients) and conditions (painting, EO, EC).

|  |  | Painting |  |  | EO |  |  | EC |  |  |
| --- | --- | --- | --- | --- | --- | --- | --- | --- | --- | --- |
|  |  | Controls<br>(n=30) | BVP<br>(n=32) | p | Controls<br>(n=30) | BVP<br>(n=34) | p | Controls<br>(n=30) | BVP<br>(n=29) | p |
| Amplitude<br>(mm) | ML | 22.9 ± 2.36 | 26.9 ± 2.22 | NS | 21.7 ± 2.36 | 29.1 ± 2.20 | NS | 25.3 ± 2.36 | 36.0 ± 2.27 | .015* |
|  | AP | 25.0 ± 2.53 | 32.0 ± 2.39 | NS | 26.3 ± 2.53 | 37.3 ± 2.36 | .020* | 31.9 ± 2.53 | 45.3 ± 2.44 | .002** |
| SD<br>(mm) | ML | 4.07 ± 0.39 | 4.79 ± 0.37 | NS | 3.86 ± 0.39 | 5.02 ± 0.36 | NS | 4.33 ± 0.39 | 5.77 ± 0.37 | NS |
|  | AP | 4.66 ± 0.45 | 5.54 ± 0.42 | NS | 5.22 ± 0.45 | 7.02 ± 0.42 | .040* | 5.53 ± 0.45 | 7.67 ± 0.43 | .008** |
| Velocity<br>(mm/s) | ML | 7.30 ± 0.74 | 8.15 ± 0.71 | NS | 6.88 ± 0.74 | 9.47 ± 0.69 | NS | 9.18 ± 0.74 | 11.54 ± 0.72 | NS |
|  | AP | 8.35 ± 0.77 | 9.84 ± 0.73 | NS | 7.87 ± 0.77 | 10.65 ± 0.72 | NS | 12.65 ± 0.77 | 18.14 ± 0.75 | NS |

BVP: bilateral vestibulopathy; EO: eyes open; EC: eyes closed; CoP: center of pressure; ML: mediolateral; AP: anteroposterior; SD: standard deviation; NS: Non-significant; \*: $p < 0.05$ , \*\*: $p < 0.01$ , \*\*\*: $p < 0.001$ .

### Mediolateral evaluation

For ML-CoP-amplitude (Fig 1A), a Group × Condition (Painting vs. EC) interaction was significant, β = 6.66, t = 2.80, p = .006. Post-hoc tests showed that patients had a greater ML-CoP-amplitude than the Control group in EC eyes closed condition (p = .015). Also, patients had greater ML-CoP-amplitude in the EC condition compared to the Painting (p < .001) and EO conditions (p < .001). The main effect of Group and the main effect of Condition were not significant.

**Fig 1.**
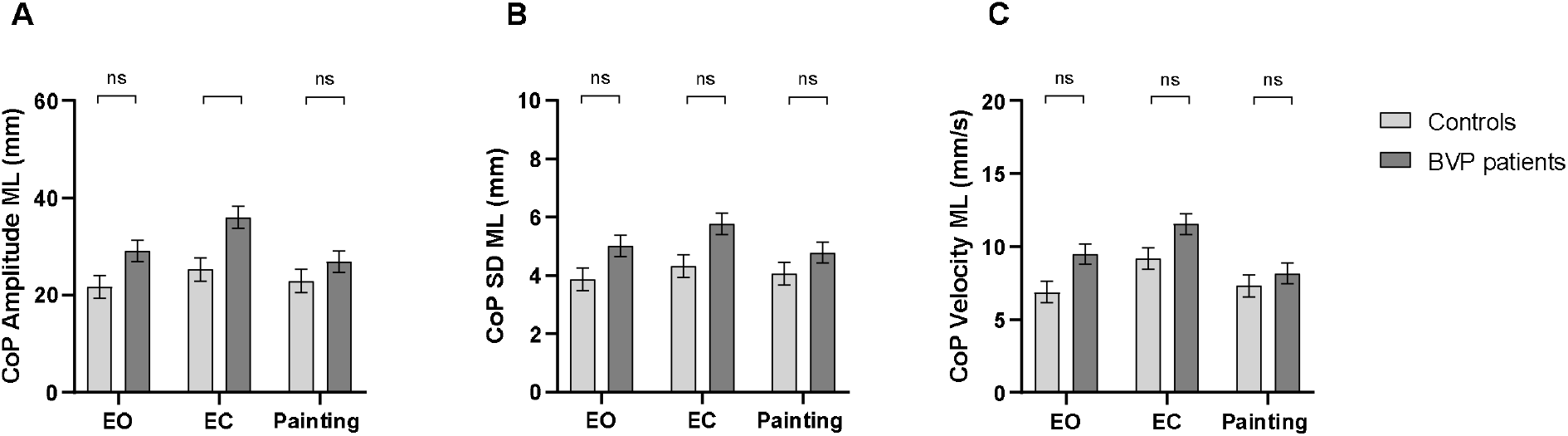
CoP mediolateral parameters of patients with BVP and controls according to sex, foot length, height and study level (marginal mean+/-SE). Comparison of the CoP amplitudes between 3 conditions (EO, EC, Painting). A. CoP amplitude; B. CoP standard deviation; C. CoP Velocity. BVP: bilateral vestibulopathy; CoP: Center of Pressure; ML: mediolateral; SD: standard deviation; EO: eyes open; EC: eyes closed.

For ML-CoP-SD (Fig 1B), no significant effects of Condition, Group, or Group × Condition interaction were found. For ML-CoP-velocity (Fig 1C), the main effect of Condition was significant (Painting vs. EC), β = 1.87, t = 2.60, p = .010. ML-CoP-velocity was greater with the eyes closed than when viewing the Painting. The main effect of Group and the Group × Condition interaction were not significant.

### Anteroposterior evaluation

For AP-CoP-amplitude (Fig 2A), the main effect of Condition was significant (Painting vs. EC), β = 6.93, t = 3.55, p = .001. AP-CoP-amplitude was higher in EC compared to the Painting condition. The main effect of Group was significant, β = 7.03, t = 2.00, p = .048. Patients had higher AP-CoP-amplitude than the Control group. A Group × Condition (Painting vs. EC) interaction was significant, β = 6.35, t = 2.29, p = .024. Post-hoc tests indicated that AP-CoP-amplitude was higher in EC compared to in EO and Painting conditions for both patients (p < .001, and p < .001, respectively) and the Control group (p =.048, and p =.005, respectively). Moreover, patients had higher AP-CoP-amplitude in both EO (p = .020) and EC (p = .002) conditions compared to the control group, but not in the Painting condition.

**Fig 2.**
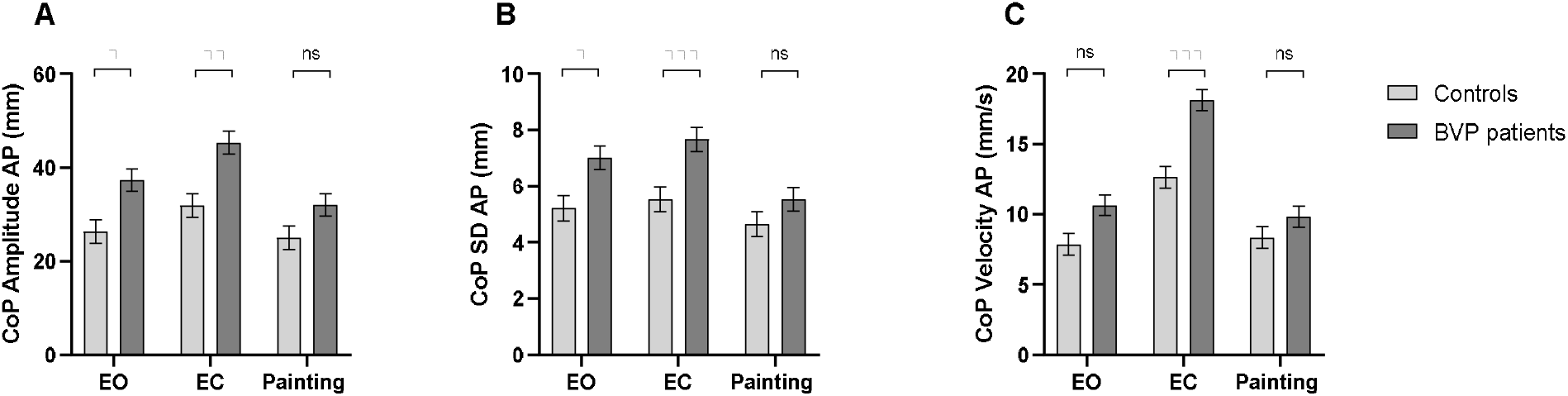
CoP anteroposterior parameters of patients with BVP and controls according to sex, foot length, height and study level (marginal mean+/-SE). Comparison of the CoP amplitudes between 3 conditions (EO, EC, Painting). A. CoP amplitude; B. CoP standard deviation; C. CoP Velocity. BVP: bilateral vestibulopathy; CoP: Center of Pressure; ML: mediolateral; SD: standard deviation; EO: eyes open; EC: eyes closed.

For AP-CoP-SD (Fig 2B), the main effect of Condition was significant (Painting vs. EC), β = 0.87, t = 2.39, p = .019. AP-CoP-SD was greater in EC compared to the Painting condition. The Group × Condition (Painting vs. EC) interaction was significant, β = 1.25, t = 2.43, p =.017. Post-hoc tests revealed that, in patients, AP-CoP-SD was greater in the EC compared to in the EO condition (p < .001), and greater in the EO compared to the Painting condition (p < .001). Moreover, patients had greater AP-CoP-SD than the Control group in EO (p = .040) and EC conditions (p = .008) but not in the Painting condition. The main effect of Group was not significant.

For AP-CoP-velocity (Fig 2C), the main effect of Condition was significant (Painting vs. EC), β = 4.30, t = 6.95, p < .001. AP-CoP-velocity was greater in EC compared to the Painting condition. The Group × Condition (Painting vs. EC) interaction was significant, β = 4.00, t = 4.55, p < .001. Post-hoc tests showed that, in the Control group, AP-CoP-velocity was higher in EC compared to in EO (p < .001) and the Painting condition (p < .001). For patients, AP-CoP-velocity was higher in EC compared to the Painting condition (p < .001). Moreover, patients had greater AP-CoP-velocity than the Control group, only in EC condition (p < .001). The main effect of Group was not significant.

## Discussion

This study aimed to examine the effects of viewing a painting on postural activity in individuals with BVP compared to healthy controls. We examined body sway while participants stood with eyes closed (EC), with eyes open while viewing a fixation cross (EO), and eyes open while viewing a painting (Painting). Our results replicated the common finding that BVP is associated with altered postural activity in both the mediolateral and anterior-posterior axes (3). Our results also replicated previous studies that have demonstrated an effect of artistic paintings on standing posture (13,14). Finally, we found that postural responses to viewing artistic paintings differed between BVP patients and healthy controls. We discuss these results in turn.

### General effects of BVP

Previous research has shown that standing body sway in persons with BVP differ from healthy controls. When standing on a flat stationary surface, Baloh et al. (1998) (3) reported that BVP patients exhibited greater amplitude and velocity of sway (compared to healthy controls). In the mediolateral axis, this effect was observed only when the eyes were open, while in the anterior-posterior axis, the effect was observed both with eyes open and with eyes closed. In the present study, a similar effect was observed only for sway amplitude in the mediolateral axis, and only when the eyes were closed. By contrast, in the anteroposterior axis, we found a significant main effect of Groups for sway amplitude. In other words, sway amplitude was greater for BVP patients, both when the eyes were closed and when they were open.

### Effects of artistic paintings on healthy controls

Cox and Klaveren (2024 and 2025) (14,15) examined the spatial dynamics of sway by analyzing the mean position of the CoP (i.e., lean) and its positional variability. Their studies compared sway during the viewing of paintings by two 20^th^ century artists, but did not include a control condition without artwork viewing. As a result, their findings are not directly comparable to ours. Kapoula and Gaertner (2015) (13) assessed sway during the viewing of artistic paintings in comparison to a control condition where participants viewed a fixation cross. They found that the velocity of CoP displacements was significantly greater during viewing of 20^th^ century paintings than when viewing the fixation cross. In contrast, in our healthy control group, we did no observe any statistically significant differences in sway between viewing the painting and viewing the fixation cross. In this sense, our results do not replicate those reported by Kapoula and Gaertner. This discrepancy may be related to differences in exposure time (60 s vs. 25.6 s) or to the nature of the paintings. Although both studies used paintings by Monet, theirs featured a standing figure with strong implied motion, while ours used a tranquil landscape with strong perspective cues. However, we found statistically significant differences in sway among healthy controls when comparing the viewing of the painting to standing with eyes closed. Nevertheless, these effects are not relevant to the question of whether artistic paintings influence postural control.

### Effects of artistic paintings on BVP patients

For BVP patients, we observed several statistically significant differences in sway between viewing the painting and standing with eyes closed (EC condition).

Notably, for BVP patients, the positional variability of the CoP in the AP axis was greater when viewing the fixation cross (EO condition) than when viewing a painting, for amplitude and standard deviation. This effect provides direct evidence on a differential influence of the artistic painting on postural control, and represents the main result of our study. It is particularly interesting to note that sway was influenced by looking at the painting (relative to the fixation cross) in BVP patients but not in healthy controls. This contrast suggests that BVP patients may exhibit sensitivity or reactivity to subtle variations in visual stimuli -in this case, the differences between a painting and a fixation cross. As mentioned, many studies have demonstrated that postural sway is systematically influenced by subtle variations in visual targets and visual tasks in healthy adults and children (7,17,18). Our findings suggest that similar effects may also occur in BVP patients, and that these effects could be more pronounced for patients than for healthy controls. This observation highlights the need for further research on how variations in visual task influences postural control in BVP patients.

## Conclusion

In our study, we measured balance in BVP patients and healthy controls when standing with their eyes closed, while viewing at a fixation cross, and an artistic painting. We evaluated several measures of postural kinematics. Our results replicated previous studies with regard to general differences in sway between patients and healthy controls, but these results didn’t replicate previous reports that looking at artistic paintings influenced the sway of healthy adults. However, we found that the standard deviation of CoP positions in the AP axis was greater when BVP patients looked at a fixation cross than when they looked an artistic painting. Also, comparing BVP patients to healthy controls, we found that the standard deviation and the amplitude in the AP axis was greater when viewing a fixation cross, and not when viewing a painting. This novel finding suggests the BVP patients retain the ability to adjust the fine details of their postural control in response to variations in visual tasks performed during posture.

## Data Availability

All data produced in the present study are available upon reasonable request to the authors.

## Acknowledgments

The authors thank the Association Française des Vestibulopathies Bilatérales for its collaboration, including all patients who participated in the study.

