## Supplementary Table for "Painting observation changes balance in patients with bilateral vestibulopathy"

|  |  | Group effect (BVP vs controls) | | | | Painting vs EO | | | | Painting vs EC | | | | Group*(Painting vs EO) | | | | Group*(Painting vs EC) | | | |
| --- | --- | --- | --- | --- | --- | --- | --- | --- | --- | --- | --- | --- | --- | --- | --- | --- | --- | --- | --- | --- | --- |
|  |  | β | *se* | t | p | β | *se* | t | p | β | *se* | t | p | β | *se* | t | p | β | *se* | t | p |
| Amplitude (mm) | ML | 4.02 | 3.27 | 1.23 | 0.221 | -1.20 | 1.67 | -0.72 | 0.475 | 2.36 | 1.672 | 1.41 | 0.161 | 3.32 | 2.32 | 1.43 | 0.155 | 6.66 | 2.38 | 2.80 | **0.006**** |
|  | AP | 7.03 | 3.51 | 2.00 | **0.048*** | 1.34 | 1.95 | 0.68 | 0.493 | 6.93 | 1.95 | 3.55 | **0.001**** | 3.97 | 2.70 | 1.47 | 0.144 | 6.35 | 2.77 | 2.29 | **0.024*** |
| SD  (mm) | ML | 0.73 | 0.54 | 1.34 | 0.181 | -0.21 | 0.26 | -0.81 | 0.419 | 0.26 | 0.26 | 0.99 | 0.324 | 0.44 | 0.36 | 1.22 | 0.226 | 0.71 | 0.37 | 1.92 | 0.058 |
|  | AP | 0.88 | 0.62 | 1.42 | 0.158 | 0.56 | 0.36 | 1.53 | 0.128 | 0.87 | 0.36 | 2.39 | **0.019*** | 0.92 | 0.50 | 1.82 | 0.070 | 1.25 | 0.52 | 2.43 | **0.017*** |
| Velocity (mm/s) | ML | 0.85 | 1.03 | 0.83 | 0.410 | -0.42 | 0.71 | -0.59 | 0.556 | 1.87 | 0.72 | 2.60 | **0.010*** | 1.74 | 0.99 | 1.75 | 0.082 | 1.52 | 1.02 | 1.49 | 0.140 |
|  | AP | 1.49 | 1.07 | 1.39 | 0.165 | -0.48 | 0.61 | -0.78 | 0.436 | 4.30 | 0.62 | 6.95 | **<0.001***** | 1.29 | 0.85 | 1.51 | 0.134 | 4.00 | 0.88 | 4.55 | **<0.001***** |

Supplementary Table. Group, Condition and Group x Condition effects on postural parameters.

*β: regression coefficient; se= standard error; t: t-value; p: p-value; BVP: bilateral vestibulopathy; EO: eyes open; EC: eyes closed; CoP: center of pressure; ML: mediolateral; AP: anteroposterior; SD: standard deviation ; * : p<0.05; ** : p<0.01; *** : p<0.001*
